# Comparative Success and Survival of Preformed Metal Crown Treatment Between the Hall Technique and Conventional Technique: An Umbrella Review

**DOI:** 10.1101/2025.05.18.25327863

**Authors:** David Okuji, Amanda Tran, Jordan Caracci, Binh Phan, Leesa Jeffries

## Abstract

**Objectives:** This umbrella review assessed systematic reviews and meta-analyses comparing the success and survival rates of Hall versus conventional techniques for primary teeth restored with preformed metal crowns.

**Methods:** A comprehensive literature search identified eligible reviews using PRISMA guidelines. Reviews were qualitatively assessed with AMSTAR 2 and quantitatively analyzed using RevMan meta-analysis. Certainty of evidence was evaluated with GRADE methodology.

**Results:** Of 116 articles screened, two reviews met inclusion criteria for quantitative analysis. Both had moderate AMSTAR 2 ratings. Meta-analysis found no significant difference between Hall and conventional techniques in pooled success and survival rates. Relative risk estimates were 0.99 (favoring conventional technique) and 1.01 (favoring Hall technique), with zero percent heterogeneity and low risk for publication bias. The GRADE assessment indicated moderate confidence, suggesting Hall technique may slightly reduce success rates while slightly increasing survival rates compared to conventional technique.

**Conclusions:** Hall and conventional techniques show comparable success and survival rates for primary molars with normal pulps or reversible pulpitis. With moderate confidence, dentists can consider Hall technique an alternative to conventional technique without additional benefit or harm related to success and survival. The Hall technique aligns with American Academy of Pediatric Dentistry guidelines as a secondary prevention method for arresting caries in children with moderate-to-high caries risk, poor cooperation, or barriers to care.

## INTRODUCTION

### Background

Oral health, in particular dental caries, has been a heavy burden on children’s health and quality of life. Children from low-income families experience significant barriers to dental care and thus often suffer from chronic caries. Untreated caries may lead to pain, localized infection, cellulitis, mental health issues, and school absences.^1^ While caries prevention is the ideal solution to this chronic disease, restorative treatment is often necessary to prevent unwanted sequelae.^1,2^

The American Academy of Pediatric Dentistry (**AAPD**) Reference Manual provides the following restorative dentistry guidance, “Traditional (conventional) stainless steel crown (**SSC)** technique (**CT**) continues to offer the advantage of full coverage to combat recurrent caries and provide strength as well as long term durability with minimal maintenance, which are desirable outcomes for caries management for high-risk children” and the “Hall Technique (**HT**) may be considered a treatment modality for carious primary molars when traditional SSC technique is not feasible due to limitations such as poor cooperation or barriers to care.^2^” The AAPD supports utilization of the HT when restoring cavitated or enlarging carious lesions in patients with a moderate- to high-risk for dental caries who exhibit poor cooperation or have other barriers to traditional restorative care.^2,3^

HT involves placing stainless steel crowns atraumatically on primary molars without the conventional tooth preparation. Current available research on Hall technique has demonstrated consistent clinical success and effectiveness.^4^ Additionally, studies have demonstrated that there is no difference in perceived pain between CT, with local anesthesia administration, and HT, without local anesthesia, which allows the dentist to offer and provide for the most effective restorative options with a personalized, patient-centered approach.^5^ Hence, factors like patient cooperation and the extent of decay should be a primary concern instead of the degree of pain that the child might experience during crown placement.

HT can also be considered a secondary prevention intervention for childhood caries. Kidd *et al*. stated, “Sealing of infected dentine within the tooth, either by a Hall crown in the primary dentition or by partial caries removal prior to placing a well-sealed filling, will also arrest the lesion.^6^” Selwitz *et al*., indicated, “Secondary prevention and treatment should focus on management of the caries process over time for individual patients, with a minimally invasive, tissue-preserving approach.^7^” Horst *et al*. further expressed, “Secondary prevention of caries requires early diagnosis and prompt treatment to reduce lesions’ complications (pain, abscess, systemic infection, etc.) and occurrence of new lesions. Secondary prevention encompasses the concept of caries lesion *arrest*, because lesions that continue to grow can cause pain, tooth loss, and may serve as a reservoir of cariogenic bacteria that can initiate new lesions.^8^” These findings, coupled with the aforementioned AAPD guidance, indicate that the HT is a secondary prevention intervention for arresting dental caries in primary molars.

However, there still lacks comprehensive studies that provide an accurate summary that address the clear and specific question: “What is the effectiveness of preformed metal crowns when comparing Hall technique and Conventional technique.” The anticipated result of this present umbrella review is that the Hall technique offers comparable effectiveness compared to Conventional technique, based on previous research.^9^

Therefore, the HT benefits societal public health and families with children with caries on primary molar teeth. Use of the treatment modality of Hall crown during the time period of the COVID-19 pandemic dramatically decreased aerosol exposure to the public and, thus, reduced the population risk of exposure to this and other air-borne pathogens.^10^ The Hall preformed metal crown (**PMC**) technique also addresses the socioeconomic and time availability needs of families with pre-cooperative children with a prevention-oriented, minimally invasive care, cost-efficient, safe, and time-efficient treatment alternative.^10^ Hence, this population of children and their families would benefit both psychologically and economically since the patients would experience more positive and less anxiety inducing appointments and the families would be able to afford more treatments, respectively.^11,12^

### Objectives

The purpose of this umbrella review was to determine if there is a difference in success and survival rates between HT and CT for children with carious primary molar teeth and a clinical diagnosis of normal pulp or reversible pulpitis. The rationale is to provide guidance to dentists with treatment alternatives for children who present with this condition. The main research question for this umbrella review is: ““In carious primary molar teeth with a clinical diagnosis of normal pulp or reversible pulpitis, is there a difference in the success and survival of PMC treatment when comparing the HT and CT?” The null hypothesis is there is no difference in PMC success or survival rates between HT and CT.

## METHODS

### Research Protocol and Registration

This umbrella review was registered on 22^nd^ April 2024 and was assigned the identification number CRD42024538335 in the PROSPERO^13^ international prospective register of systematic reviews hosted by the National Institute for Health Research, Center for Reviews and Dissemination, University of York, UK.

### Reporting format

This review was adapted and reported according to the Preferred Reporting Items for Systematic Reviews and Meta-Analysis (**PRISMA**)^14^ and the Cochrane Handbook for Systematic Reviews of Interventions^15^ throughout the process of this umbrella review.

### Eligibility criteria

#### Population, Intervention, Comparator, Outcome (*PICO)*

The population included children, regardless of sex, race, socioeconomic status, health status, or geographic location, who presented with primary molars requiring PMC treatment with diagnoses of normal pulp or reversible pulpitis. The intervention was PMC treatment using HT (i.e., cementation of an SSC over a caries-affected primary molar without local anesthetic, caries removal, or tooth preparation^2^, which seals a dentine carious lesion and allows for inactivation of carious lesion as well as the restoration of form and function^16^). The comparator was PMC treatment using CT (i.e., including non-selective caries removal to hard dentine, with use of local anesthesia^16^). The following outcomes were studied:

1. Successful treatment was defined throughout the 12- to 24-month follow-up period as 1) Symptom-free (without pain, swelling, infection, pathological mobility), 2) No radiographic signs of pathological radiolucency, 3) Satisfactory restoration with no replacement required.
2. Survival was defined, throughout the 12- to 24-month follow-up period, as the presence of the preformed metal crown without any major clinical or radiographic failure (e.g., pain, infection, crown lost and tooth unrestorable, required pulp treatment or extraction, pathological radiolucency).

#### Criteria for selecting reviews for inclusion

The reviewed studies were designed as a systematic review, with meta-analysis of randomized controlled studies, which analyzed the restoration of asymptomatic primary molars with caries into the dentin with either HT or CT. The studies must have compared HT and CT outcomes quantified as odds ratio or risk ratio for success, failure, or survival rate.

#### Exclusion Criteria

Reviewed studies, which were not designed as a systematic review with meta-analysis of randomized controlled studies, were excluded from this review. Reviews that analyzed primary carious molars with incipient enamel lesions, symptomatic molars, or molars with the diagnosis of irreversible pulpitis or necrotic were also excluded. Articles which were classified as case reports, comments, laboratory studies, letters, and narrative reviews were excluded.

### Literature Search

#### Search strategy, information sources, and selection process

An initial literature search was conducted on 3^rd^ September 2023 in all relevant publications in the PubMed, Embase, Cumulative Index to Nursing & Allied Health (**CINAHL**), Scopus, Web of Science, Cochrane Database of Systematic Reviews, Dentistry & Oral Sciences Source (**DOSS**) and, Health and Psychosocial Instruments (**HaPI**) databases, as well searching the grey literature. Hand searching and checking reference lists were also used to identify additional relevant records. The search strategy was composed of the following keywords and Boolean operators “systematic review,” “review,” or “meta-analysis,” using the keywords “Hall technique,” “conventional technique,” “early childhood caries,” “performed metal crowns,” “stainless steel crowns,” “primary molars,” and “dentin” and applied filters for “meta-analysis,” “review,” “systematic review.” An updated search was conducted on 14^th^ April 2025, to identify additional studies.

#### Study selection

For the study selection, two selected authors independently screened the titles and abstracts using an Excel spreadsheet tool (Microsoft Corporation, Redmond, WA) to eliminate duplicates and read the full text of all papers to identify relevant systematic reviews, with meta-analyses. Discrepancies with selected reviews were resolved through discussion and mutual agreement by the two researchers.

The results of the study selection outcomes were populated into a Preferred Reporting Items for Systematic Reviews and Meta-Analyses (PRISMA) flow diagram.^14^

### Data Collection and Analysis

#### Synthesis methods

The methods utilized to synthesize the data from the reviewed articles included 1) Excel spreadsheet (Microsoft Corporation, Redmond, WA) to compile the characteristics of the included studies, 2) corrected covered area (**CCA**) citation matrix analysis to measure the degree of overlap, 3) “A MeaSurement Tool to Assess systematic Reviews” (**AMSTAR**) 2 to conduct the quality appraisal, risk of bias, heterogeneity of the data, 4) RevMan 5.4 software application *(The Cochrane Collaboration Review Manager 5, 2020, Copenhagen, Denmark)* to meta-analyze the effect size of variables, heterogeneity of the data, publication bias, and 5) Grading of Recommendations, Assessment, Development and Evaluation (**GRADE**) to assess the certainty of evidence (**COE**).

#### Data collection process and data items

Qualitative data extraction for the characteristics of the included systematic review and meta-analysis studies was completed by two of this study’s authors who independently focused on collecting details on the diagnostic tests, author names, publication year, journal name, research question, search strategies, study design, outcomes, assessment tools, and conclusions for each study marked for inclusion in this investigation. Disagreements were resolved through discussion until mutual agreement was reached between the two researchers.

Quantitative data extraction was completed by the same two independent reviewers. Using RevMan 5.4 software, the meta-analysis included the author names, year of publication, number and study design of the primary studies, point estimates with confidence intervals, and heterogeneity statistics.

#### Analysis of degree of overlap

A citation matrix was generated to calculate the CCA. This analysis classifies the degree of overlap as “slight” (zero to five percent), “moderate” (six to ten percent), “high” (11 to 15 percent) or “very high” overlap (greater than 15 percent).^17^ The formula used is CCA = (N-r) / (rc-r), where “N” is number of included primary studies, “r” equals the number of index publications, and “c” includes the number of reviews.

#### Quality appraisal

The quality appraisal was conducted with the AMSTAR 2 tool, which enables the assessment of systematic reviews of randomized and non-randomized studies of healthcare interventions. The tool contains 16 domain items to assess the quality of included systematic reviews.^18^ The domain questions are designed so that a “yes” answer denotes a positive result and when information needed was present in the study. The “no” answer denotes when no information is available to rate. The “partial yes” answer denotes when partial information is provided in the article. The tool provides an overall rating based on weaknesses in critical domains.^18^

#### Risk of bias assessment

The risk of bias (**RoB**) assessment is included in two of the domain items of the AMSTAR 2 tool.^18^ The RoB from individual primary studies and from the interpretation of the results of the review are included in domain “item 9” and “item 13” shown in Figure 2. “Item 15” in Figure 2 specifically assessed the risk of publication bias. Separately with the RevMan 5.4 software, publication bias was separately analyzed with funnel-plot analytic methodology, which is based on evidence for the effect of small-studies.

#### Estimation of common effect size

The primary aim of this umbrella review was to facilitate a straightforward comparison of the effects across various factors examined by utilizing a consistent measure of effect size.

Since systematic reviews and meta-analyses incorporate different measures based on the design and analytical methods of the included studies, it was important to establish a common effect size to enable an overall comparison. Conversion of all effect size into equivalent odds or risk ratios was planned, using the RevMan 5.4 software.

#### Heterogeneity of data

The heterogeneity of the data was analyzed using the I² statistic, which is quantified with the RevMan 5.4 software. In meta-analysis, the I² statistic quantifies the proportion of variability in a meta-analysis due to differences between included trials, rather than random sampling error. It ranges from 0% to 100%, with higher values indicating greater heterogeneity. As suggested guidelines, I² values can be categorized as low (25%), moderate (50%), and high (75%) heterogeneity.^19^

#### Certainty of evidence assessment

The certainty of evidence (**COE**) for quantitative outcome data utilized the Grading of Recommendations, Assessment, Development and Evaluation (**GRADE**)^20^ methodology to assess and report the confidence the investigators have in the effect estimate for each pre-defined, clinically important outcome of interest in the umbrella review. The investigators extracted and reported the GRADE assessments presented in the included systematic reviews and meta-analyses.

## RESULTS

### Description of included reviews

#### Literature search results and study selection

The search resulted in a total of 116 references, 105 sources from databases and 11 from registers, that underwent screening. After two screening stages, a total of two systematic reviews with meta-analyses (Chua 2023^16^ and Hu 2022^21^) were included in this umbrella review. The details of the results of each search and screening stage can be found in PRISMA flow diagram (Figure 1).

**Figure 1:**
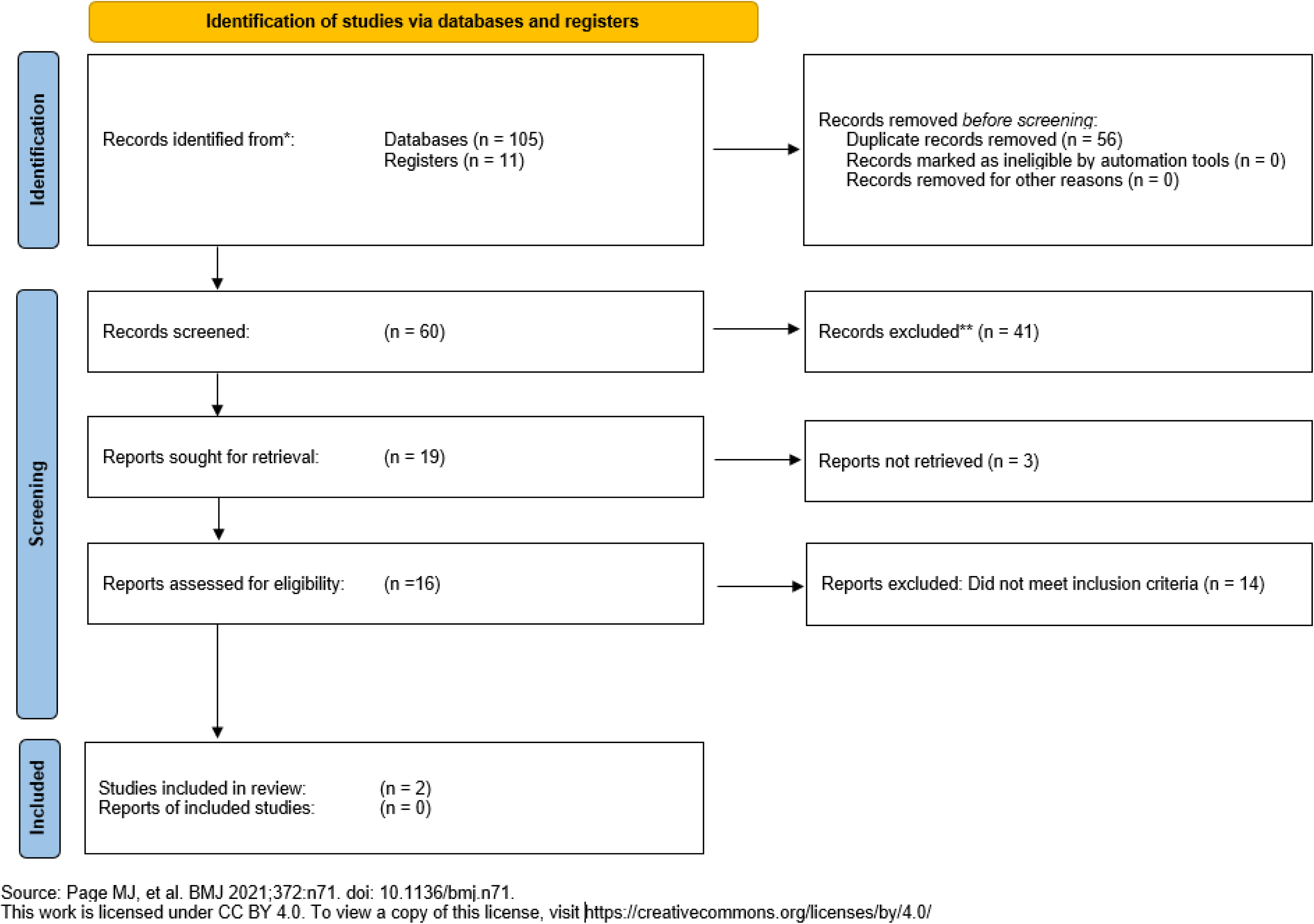
Preferred Reporting Items for Systematic reviews and Meta-Analyses (PRISMA) flow diagram of identification and selection of included reviews.

#### Study characteristics

As shown in Table 1, one review was published in 2023 (Singapore and Qatar)] and one in 2022 (Singapore, United Kingdom, and Germany).^16,21^ The number of databases searched by the included reviews ranged from five to eight, with the search period end-year ranging from 2007 to 2022. Both reviews included a search restriction for language to English. The primary design for both reviews was randomized clinical trials with respective number of studies as four and two.^16,21^ The primary research question for the two reviews were respectively: 1) When restoring carious primary molars with preformed metal crowns in children, how does the HT compare against the CT in terms of overall success and survival? ^16^” and 2) “Do patients with dentine carious lesions in primary molars that are managed with Hall technique crowns compared with conventional restoration approaches, other minimal intervention dentistry techniques and no treatment have different outcomes, in terms of treatment success and failure?^21^”

**Table 1.** Characteristics of Included Articles.

| Authors, year, journal (country) | Search strategy: Databases | Search strategy: Search period start date | Search strategy: Search period end date | Search strategy: Language restriction | Study design of the included studies | Study design of the included studies: Number of studies | Research question | Study population | Indicator/ Intervention | Comparator/ Control | Conclusions | Assessment tools | Meta-analysis performed |
| --- | --- | --- | --- | --- | --- | --- | --- | --- | --- | --- | --- | --- | --- |
| Chua *et al.*, 2023, International Journal of Paediatric Dentistry (Singapore, Qatar) | 1. Cochrane Central Register of Controlled Trials<br>2. Embase<br>3. PubMed<br>4. Scopus<br>5. Web of Science<br>6. Grey Literature:<br>a. Google Scholar<br>b. opengrey.eu | 6/30/2021 | 8/23/2022 | Restricted to English language | Randomized clinical trials | Four (4) included studies for meta-analysis were designed as randomized controlled trials. | When restoring carious primary molars with preformed metal crowns (PMCs) in children, how does the HT compare against the CT in terms of overall success and survival?' | Children aged 2–12, with dental caries affecting primary molars indicated for PMC but without prior symptoms of pulp or periapical pathology. | Placement of a crown according to the Hall technique (PMC is cemented to seal a dentine carious lesion, allowing for inactivation of carious lesion as well as the restoration of form and function). | Conventional PMC restoration (including non-selective caries removal to hard dentine). | Success and or survival of PMCs placed using HT versus CT over a minimum period of 12 months.<br>Success defined as the presence of the PMC without any major or minor failures.<br>Survival defined as the presence of the PMC without any major failures.<br>1. Greater consideration may be given towards using the HT as part of standard treatment procedures in managing carious primary molars. Future studies should standardize reporting of outcomes to facilitate a more homogeneous pool of data for future meta-analysis.<br>2. For overall success, there were no significant differences were found between HT and CT at 12 and 24 months (p > .05).<br>3. For survival, there were no significant differences found between HT and CT at 12 and 24 months (p > .05). | National Institutes of Health Quality Assessment tool | Yes |
| Hu *et al.*, 2022, Japanese Dental Science Review (Singapore, United Kingdom, Germany) | 1. Cochrane Central Register of Controlled Trials<br>2. Embase<br>3. Epistemonikos<br>4. PubMed<br>5. MEDLINE | 1/1/2007 | 8/31/2022 | Restricted to English language | Randomized clinical trials | Two (2) included studies for meta-analysis were designed as randomized controlled trials. | Do patients with dentine carious lesions in primary molars that are managed with Hall technique crowns compared with conventional restoration approaches, other minimal intervention dentistry (MID) techniques and no treatment have different outcomes, in terms of treatment success and failure? | Children with untreated carious lesion(s) extending into dentine in primary molars that required intervention to limit caries progression. Only teeth without existing restorations were considered in order to exclude the possibility of the dental pulps being compromised by previous treatments. | In Hall Technique (HT), a preformed metal crown (PMC) is cemented over a primary molar to seal a dentine carious lesion, allowing for inactivation of carious lesion as well as the restoration of form and function. | Conventional restoration approaches including non-selective caries removal to hard dentine (direct restorations, conventional PMCs), other MID techniques (Atraumatic Restorative Technique [ART], 38 % Silver Diammine Fluoride [SDF] application, Non-restorative Cavity Control [NRCC]), placebo and no treatment. | Clinical outcome: Success defined as retention of deciduous tooth and/or absence of pulpal symptoms.<br>1. Hall Technique (HT) is a successful option for the management of caries in primary teeth, particularly for proximal or multi-surface dentine lesions. It is well-tolerated by children and acceptable to parent, with mild adverse effects reported.<br>2. Specifically, HT has 1.80 greater odds of success compared to conventional technique with preformed metal crowns. | Cochrane Risk of Bias 2 (RoB2) tool | Yes |

The two reviews included the following populations, indicators, comparators, conclusions, assessment tools, and meta-analyses. Both reviews examined populations of children with caries affected primary molars with caries into dentin, without pulpal pathology, and indicated for PMC treatment; defined the indicator as Hall PMC technique allowing for inactivation of carious lesion as well as the restoration of form and function; and the comparator as conventional PMC technique with non-selective caries removal to hard dentin. The conclusions for the two reviews were aligned with Chua *et al*. concluding that there were no significant differences between HT and CT for overall success and failure and Hu *et al*. concluding that HT has 1.80 greater odds of success compared to conventional technique with preformed metal crowns.^16,21^ For quality assessment tools, Chua *et al*. utilized the National Institutes of Health Quality Assessment tool and Hu *et al*. utilized the Cochrane Risk of Bias 2 tool.^16,21^ Both reviews conducted meta-analyses.

#### Excluded studies

A total of 12 studies that underwent full-text screening were excluded from this umbrella review because they did not fulfill the inclusion and selection criteria.

### Data Analysis and Results of Synthesis

#### Overlap of studies

Table 2 shows the results of the CCA analysis. With two reviews meeting the inclusion criteria, including a total of four index studies which focused on success and/or survival rate of HT compared to CT, the degree of overlap between the reviewed studies was 50 percent.

**Table 2.**
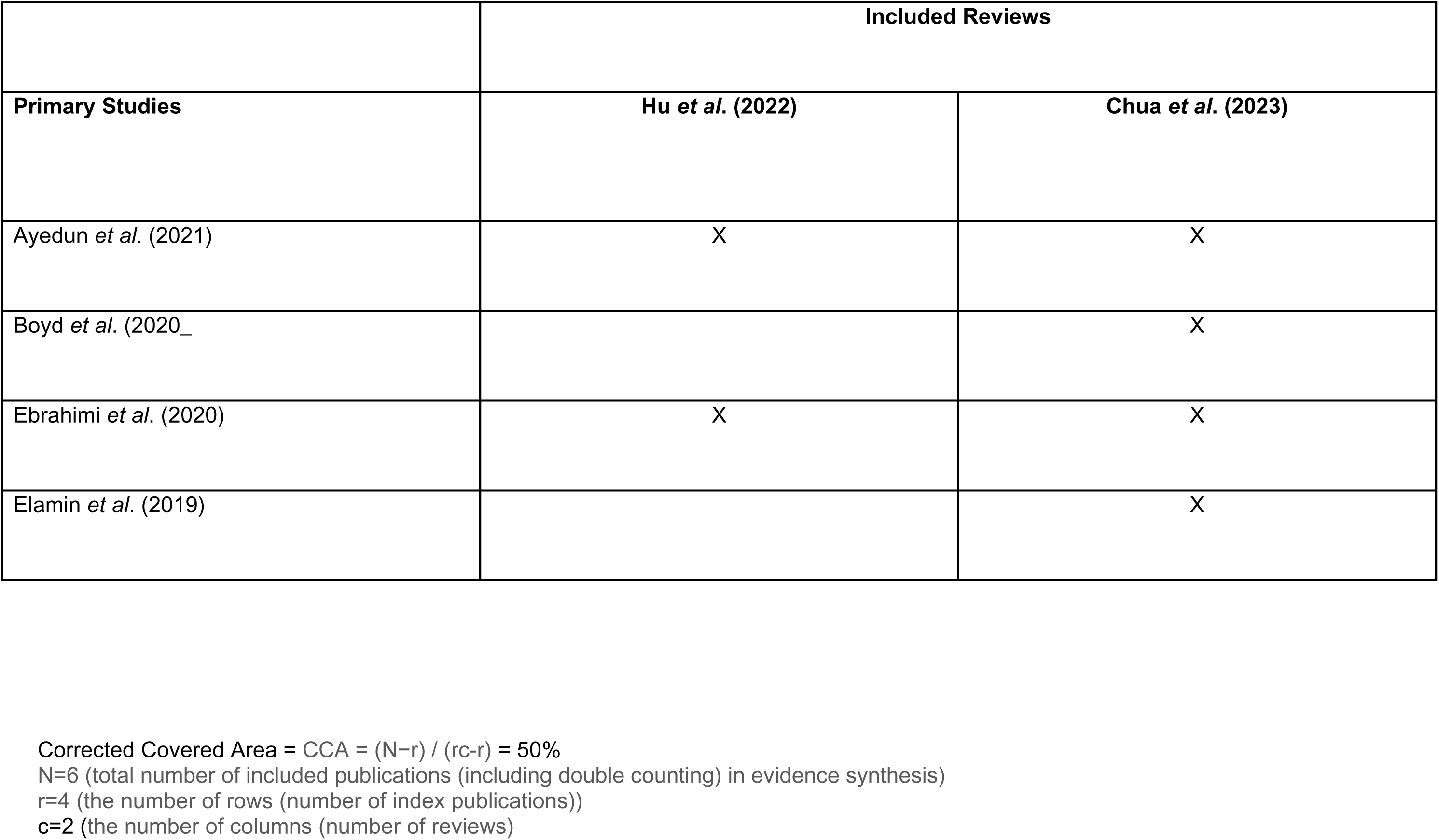
Corrected Covered Area (CCA)

|  | Included Reviews |  |
| --- | --- | --- |
| Primary Studies | Hu *et al.* (2022) | Chua *et al.* (2023) |
| Ayedun *et al.* (2021) | X | X |
| Boyd *et al.* (2020_ |  | X |
| Ebrahimi *et al.* (2020) | X | X |
| Elamin *et al.* (2019) |  | X |

Corrected Covered Area = CCA = (N-r) / (rc-r) = 50% N=6 (total number of included publications (including double counting) in evidence synthesis) r=4 (the number of rows (number of index publications)) c=2 (the number of columns (number of reviews))

#### Methodological quality of included reviews

##### Quality appraisal

The results of the AMSTAR 2 quality appraisal of the two reviews are presented in Figure 2. Two reviews^16,21^ (100 percent) were classified as being of moderate quality per the AMSTAR 2 rating scale. Both reviews^16,21^ (100 percent) partially conducted the critical investigation of publication bias (item 15), which contributed to downgrading the overall quality assessments for both reviews to the moderate classification.

**Figure 2.**
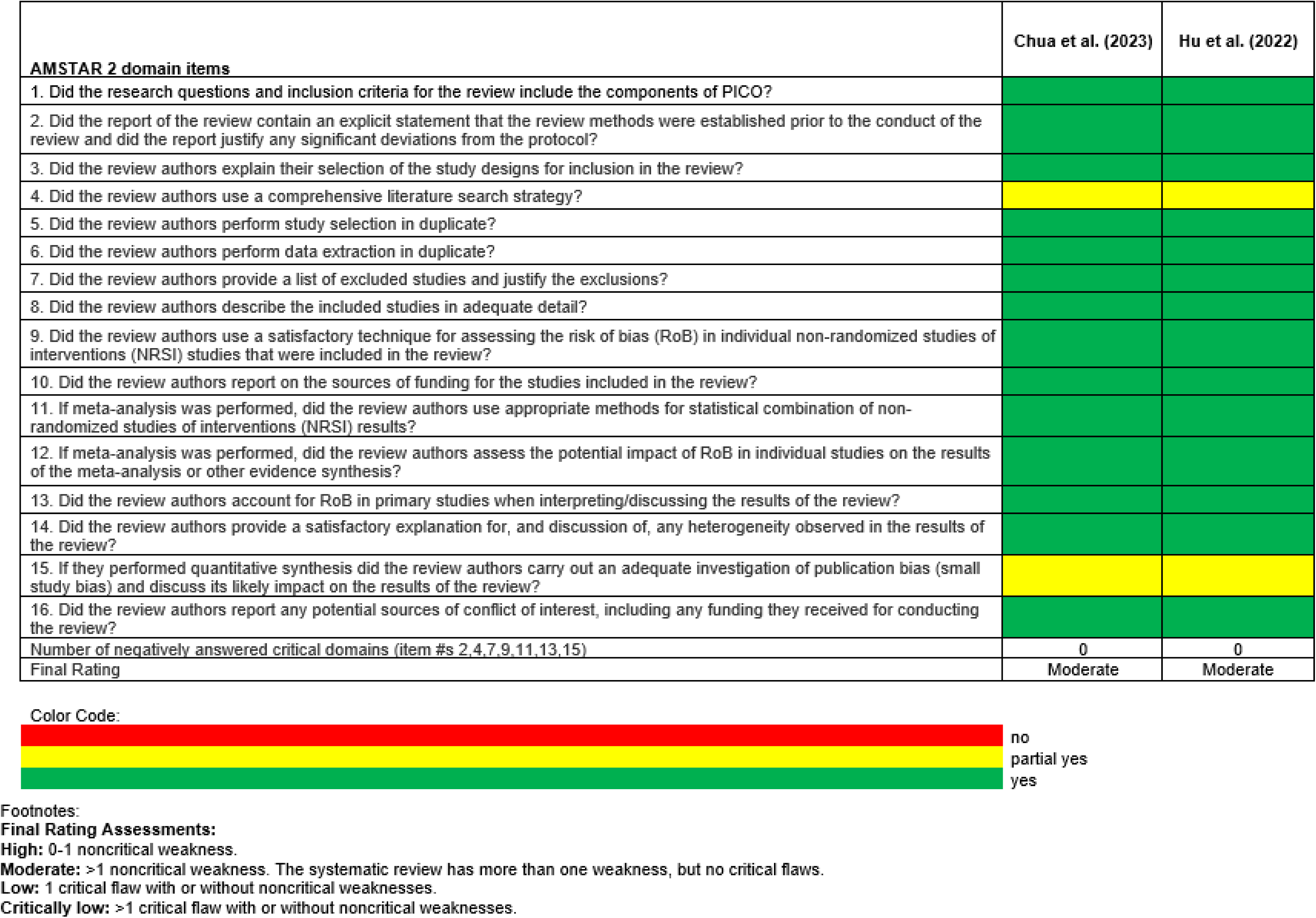
AMSTAR 2 assessment.

##### Risk of Bias (ROB)

AMSTAR 2 was used to assess quality appraisal and the risk of bias. The results are depicted in Figure 2. The results show two (100 percent) of the included reviews had “yes” responses for “item 9” and “item 13” in the AMSTAR 2 assessment. Item 9 asks, *“*Did the review authors use a satisfactory technique for assessing the RoB in individual studies that were included in the review?” *and item 13, “*Did the review authors account for RoB in primary studies when interpreting/discussing the results of the review? The risk of bias assessment tools utilized by the two reviews included the National Institutes of Health Quality Assessment tool (Chua *et al*., 2022), and Cochrane Risk of Bias 2 (RoB2) tool (Hu *et al*., 2022) and were found to be satisfactorily addressed.

##### Publication Bias

From the AMSTAR 2 tool (Figure 2), both (100 percent) of the included reviews had “partial yes” responses for “item 15” in the AMSTAR 2 assessment which asked, *“*If they performed quantitative synthesis did the review authors carry out an adequate investigation of publication bias (small study bias) and discuss its likely impact on the results of the review?” These responses contributed to lowering the final quality appraisal ratings for both included reviews to the moderate level.

Although the quantitative meta-analysis for both included reviews only totaled five primary databases, the funnel-plot analyses showed that the likelihood of publication bias was low for both success rate (Figure 3.b.) and survival rate (Figure 3.d.) because of the symmetry and homogeneity along the center axis with the relative risk nearly equal to 1.00.^22^

**Figure 3.a.**
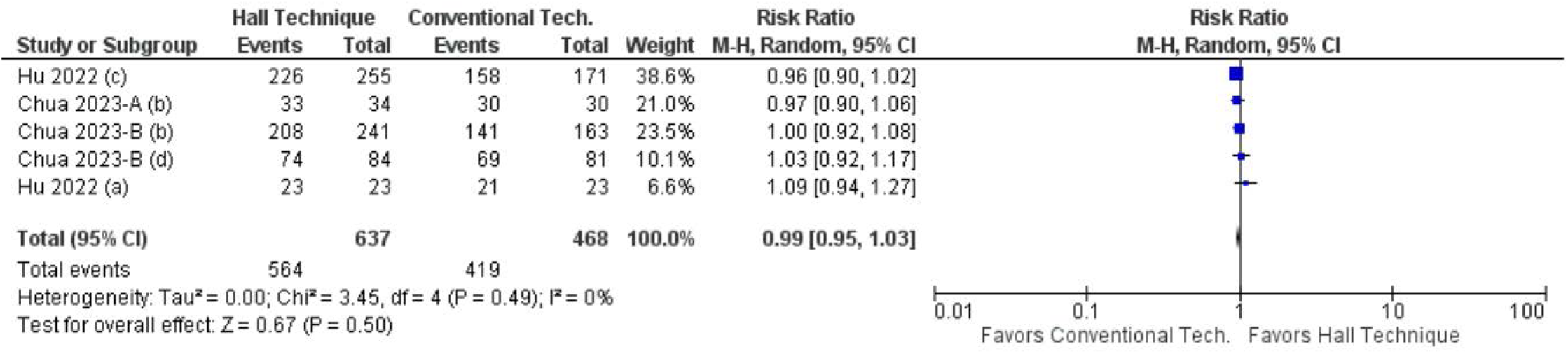
Forest plot of comparison. Success Rate: Hall versus Conventional Crown Technique.

**Figure 3.b.**
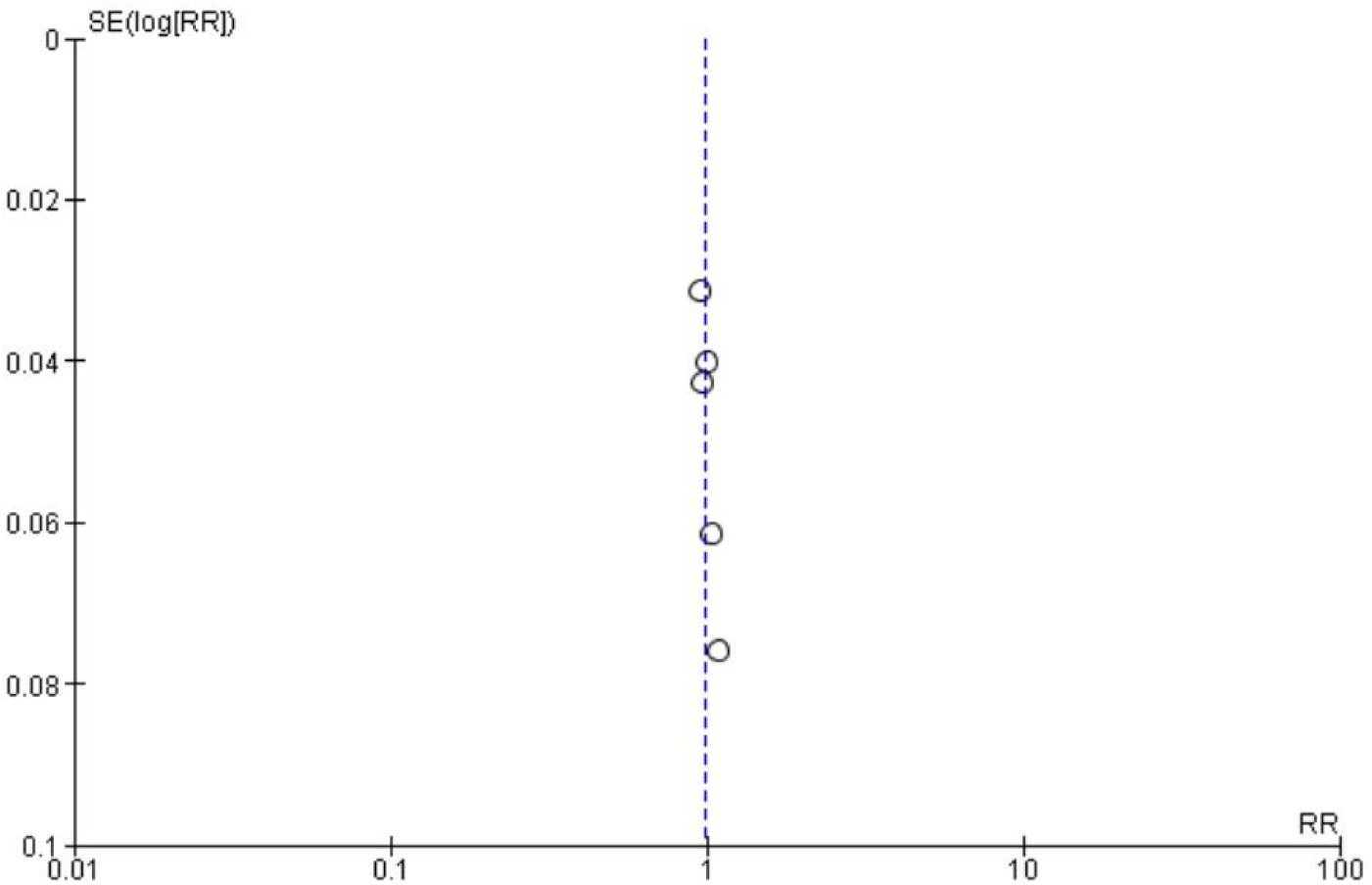
Funnel plot of comparison. Success Rate: Hall versus Conventional Crown Technique.

**Figure 3.c.:**
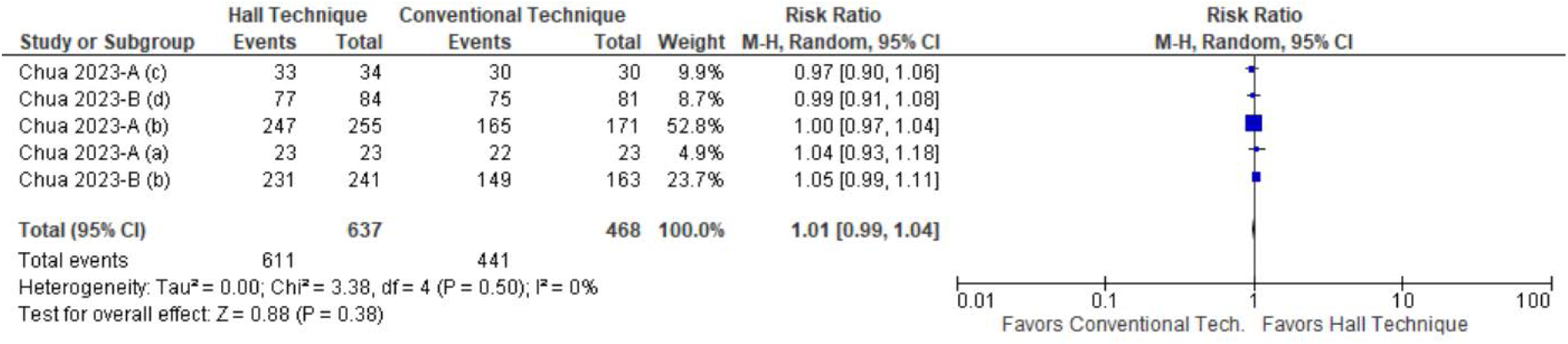
Forest plot of comparison. Survival Rate: Hall vs Conventional Crown Technique.

**Figure 3.d.**
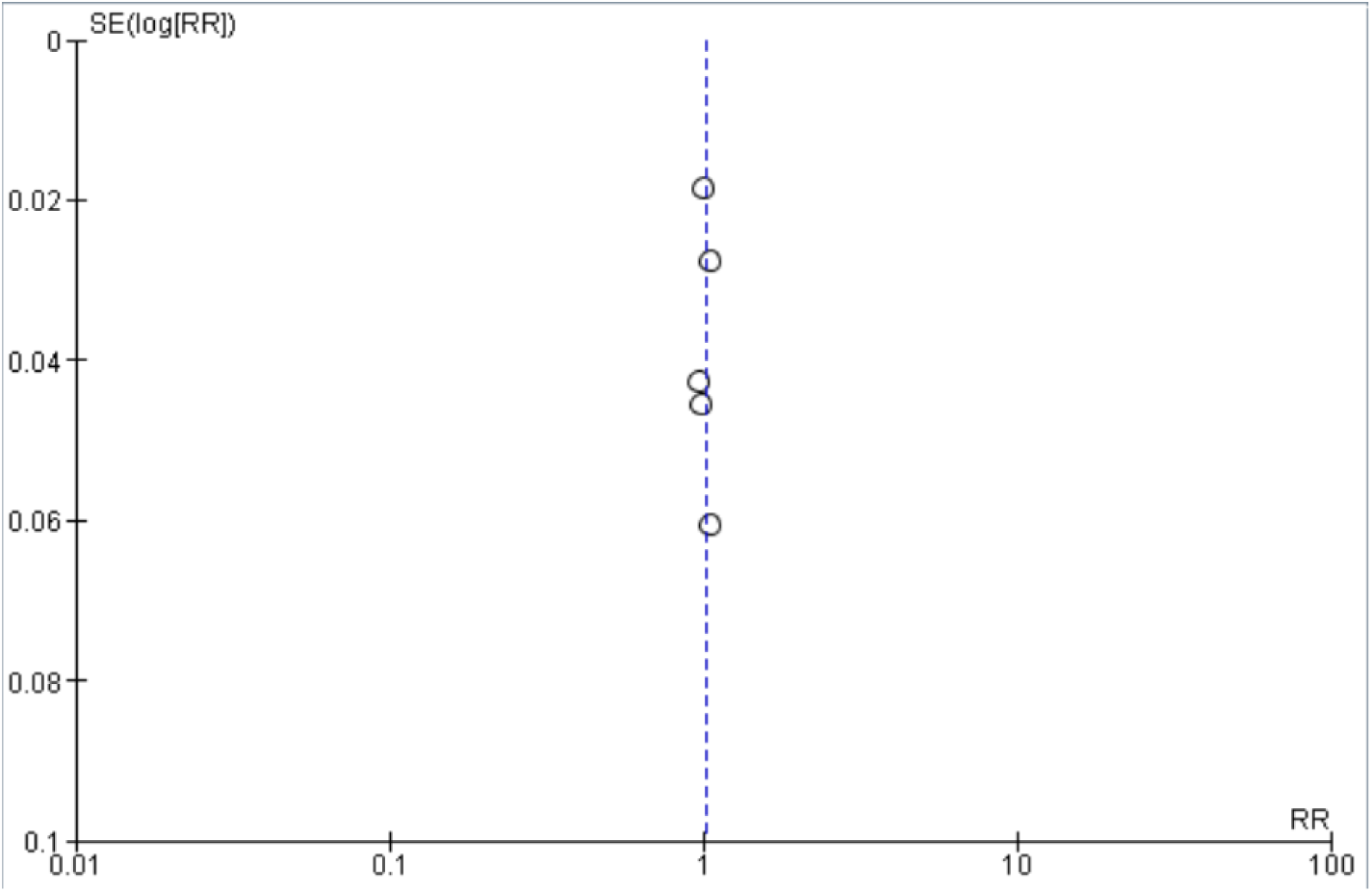
Funnel plot of comparison. Survival Rate: Hall vs Conventional Crown Technique.

##### Effects of interventions

Summaries for the meta-analyses of the success rates of HT versus CT PMCs and survival rates of HT versus CT PMCs are visualized in Figures 3.a. and 3.c., respectively.

The analysis for success rates of HT versus CT PMCs included data from a total of five randomized controlled trials for primary study subgroup analysis, three from the Chua *et al*. (2023-A (b),^23^, -B(b),^23^ -B(d)^24^) data and two from the Hu *et al*. (2022 (a)^25^ and (c)^26^) data. The meta-analysis demonstrated an overall pooled effect with the risk ratio at 0.99, confidence interval of 0.95 to 1.03, and Z-test for overall effect of 0.67 at P = 0.50. Hence, although the success rate slightly favors CT, it is not statistically significant.

The analysis for survival rates of HT versus CT PMCs included data from a total of five randomized controlled trials for primary study subgroup analysis from the Chua *et al*. (2023-A (a),^25^ (b),^23^ (c),^26^ -B(b),^23^ (d)^24^) data. The meta-analysis demonstrated an overall effect for a risk ratio of 1.01, confidence interval of 0.99 to 1.04, and Z-test for overall effect of 0.88 at P = 0.38. Although the survival rate slightly favors HT, it is not statistically significant.

##### Heterogeneity of Data

Summaries for the meta-analyses of the data heterogeneity of HT versus CT PMC success and survival rates are respectively visualized in Figures 3.a. and 3.c.

For success rate of HT versus CT PMCs, the test for heterogeneity I² statistic was calculated as zero percent at P = 0.49. For survival rate of HT versus CT PMCs, the I² statistic was calculated as zero percent at P = 0.50.

Additionally, from the AMSTAR 2 assessment (Figure 2), both included reviews had “yes” responses for “item 14” in the AMSTAR 2 assessment, which asked, “Did the review authors provide a satisfactory explanation for, and discussion of, any heterogeneity observed in the results of the review?”

Therefore, for both success and survival rates of HT versus CT, the heterogeneity is categorized as low.

##### Certainty of Evidence

Table 3 shows the GRADE summary of findings with the overall certainty of evidence ratings and comments for comparing success and survival rates for HT versus CT treatment in primary molars.

**Table 3.**
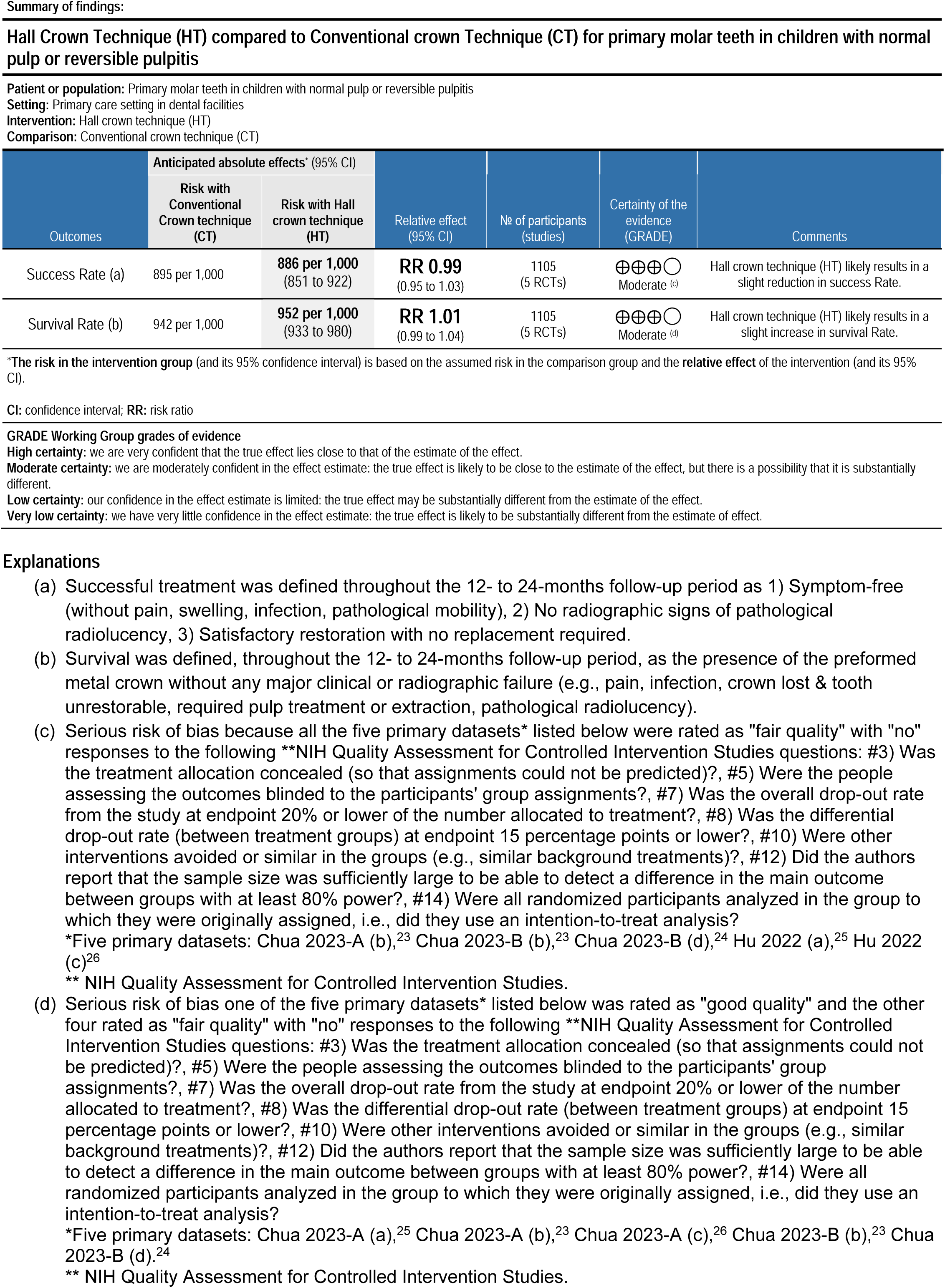
GRADE PRO Summary of findings.

| Outcomes | Anticipated absolute effects* (95% CI) |  | Relative effect (95% CI) | No of participants (studies) | Certainty of the evidence (GRADE) | Comments |
| --- | --- | --- | --- | --- | --- | --- |
|  | Risk with Conventional Crown technique (CT) | Risk with Hall crown technique (HT) |  |  |  |  |
| Success Rate (a) | 895 per 1,000 | 886 per 1,000 (851 to 922) | RR 0.99 (0.95 to 1.03) | 1105 (5 RCTs) | ⊕⊕⊕○ Moderate (c) | Hall crown technique (HT) likely results in a slight reduction in success Rate. |
| Survival Rate (b) | 942 per 1,000 | 952 per 1,000 (933 to 980) | RR 1.01 (0.99 to 1.04) | 1105 (5 RCTs) | ⊕⊕⊕○ Moderate (d) | Hall crown technique (HT) likely results in a slight increase in survival Rate. |

\*The risk in the intervention group (and its 95% confidence interval) is based on the assumed risk in the comparison group and the relative effect of the intervention (and its 95% CI).
CI: confidence interval; RR: risk ratio
GRADE Working Group grades of evidence
High certainty: we are very confident that the true effect lies close to that of the estimate of the effect. Moderate certainty: we are moderately confident in the effect estimate: the true effect is likely to be close to the estimate of the effect, but there is a possibility that it is substantially different. Low certainty: our confidence in the effect estimate is limited: the true effect may be substantially different from the estimate of the effect. Very low certainty: we have very little confidence in the effect estimate: the true effect is likely to be substantially different from the estimate of effect.
Explanations

For success rate, HT intervention yielded a risk of 886 success per 1,000 events, with an average relative risk of 0.99 compared to CT. The GRADE certainty of evidence was rated as “moderate” confidence with the overarching comment, “HT likely results in a slight reduction in success rate” compared to CT.

For survival rate, HT intervention yielded a risk of 952 per 1,000 events, with an average relative risk of 1.01 compared to CT. The GRADE certainty of evidence was rated as “moderate” confidence with the overarching comment, “HT likely results in a slight increase in survival rate” compared to CT.

## DISCUSSION

### Summary of the main results

The main results from this umbrella review found that with moderate confidence the success and survival rates between the Hall and Conventional techniques for PMC restorations were essentially equal for HT and CT without statistical significance, with CT having a slight increase in success rate and HT a slight increase in survival rate. Hence, the findings support the null hypothesis is there is no difference in PMC success or survival rates between HT and CT.

### Overall completeness and applicability of evidence

The included reviews in this umbrella review provided information regarding two specific outcomes of interest, success and survival rates between HT and CT. The included reviews incorporated a broad sample of participants who were treated in different settings. This, along with findings that were similar among the randomized controlled trials demonstrate the generalizability of the evidence to a broad range of populations.

### Quality of the evidence

Based upon the AMSTAR 2 assessment, meta-analysis findings, and the GRADE assessment, the overall quality of the evidence is presented with moderate confidence.

The AMSTAR 2 assessment showed moderate quality ratings for the two included reviews. The meta-analysis findings demonstrated nearly equal mean effects between HT and CT without statistical significance, along with low heterogeneity of the data and low risk of publication bias. The GRADE summary of findings provided certainty of evidence with moderate confidence, which supports the null hypothesis that there is no difference between success and survival rates between HT and CT.

### Potential biases in the overview process

This umbrella review adapted the steps according to the Preferred Reporting Items for Systematic Reviews and Meta-Analysis and the Cochrane Handbook for Systematic Reviews of Interventions. Potential bias was mitigated with funnel plot analysis to assess the risk of publication bias and subgroup analysis which showed low statistical heterogeneity.

The strengths of this umbrella review include the performance of a systematic and comprehensive search strategy in multiple electronic databases to avoid missing relevant systematic reviews, with meta-analyses, and utilization of two reviewers independently assessed the quality of evidence. The AMSTAR 2 assessment was used as a critical assessment tool for systematic reviews in this umbrella review. Another strength was the extraction of subgroup data from seven primary studies with nearly 1,105 subjects. Finally, utilization of the GRADE tool with its summary of findings supports the certainty of evidence which was deemed as moderate.

Limitations of this umbrella review included the high CCA value where 50 percent of index reviews appeared multiple times across the two included reviews, which may have contributed to the weighting of the results and was mitigated by the utilization of subgroup analysis. Despite this limitation, this review provides a comprehensive synthesis of the evidence on the success and survival rates between HT and CT. Dentists who treat children can be more confident that using HT in place of CT will be very unlikely to cause any added benefit or harm to children related to PMC success and survivability.

### Agreements and disagreements with other studies and/or reviews

A significant area of agreement among the analyzed systematic reviews is the comparable short-term success and survival rates of HT and CT for restoring carious primary molars in children. Individual studies by Binladen *et al.*^27^ and Elamin *et al.*^24^ reported high success and survival rates for both techniques within the first 12 to 24 months, with no statistically significant differences observed between them. Similarly, Ludwig *et al.*^28^ found similar success rates for stainless steel crowns placed using either technique. This convergence of evidence suggests that both techniques are effective in the short to medium term for managing caries in primary molars.

There are some areas of disagreement and conflicting findings among the systematic reviews. One such area pertains to the potential differences in long-term success and survival rates between the two techniques. While the previously referenced reviews offer limited data on direct comparisons beyond 24 months, the study by Şen Yavuz *et al.*^29^ showing similar 60-month survival between Hall Technique PMCs and conventional compomer restorations hints at the potential for long-term effectiveness of the Hall Technique, although a direct comparison with conventional PMCs over this extended period is lacking in the provided literature.

Another point of divergence is the assertion of superiority of one technique over the other. Badar *et al.*^4^ concluded that their meta-analysis significantly favored the Hall Technique in terms of success. This discrepancy could be due to the inclusion of different primary studies in each review, variations in the definition of “conventional restorations” (which might have included direct fillings in some cases), or differences in the statistical methods employed for the meta-analysis. The explicit statement by Badar *et al.*^4^ that the Hall Technique significantly outperformed conventional restorations contrasts with the more neutral findings of other reviews, indicating a lack of complete consensus on this matter.

Furthermore, the methodological differences and potential biases within the primary studies included in the systematic reviews may contribute to the variability in findings. Tedesco *et al.*^30^ noted that the primary studies which were analyzed had a range of risk of bias from low to high, which could affect the reliability of the pooled success rates. These acknowledged limitations in the quality and methodology of some primary studies warrant caution when interpreting the pooled results and highlight the need for ongoing research with rigorous designs.

### Implications for research

To further refine the understanding of the comparative effectiveness of the Hall Crown and Conventional Crown techniques, there is a significant need for more high-quality research. Specifically, future studies should prioritize conducting randomized controlled trials that directly compare the two techniques with long-term follow-up periods extending beyond 24 months. This will provide more definitive evidence on any potential differences in success and survival rates over the lifespan of the primary molars.

Future research should also focus on standardizing the reporting of outcomes. This includes the use of clearly defined and universally accepted success and failure criteria to facilitate more robust and meaningful comparisons across different studies and to enable more reliable meta-analyses in the future.^16^

Investigating the cost-effectiveness of both techniques in various clinical settings is also crucial for informing healthcare policy and resource allocation.^16^ Studies should explore the direct and indirect costs associated with each technique, including treatment time, materials, and the need for retreatment.

The impact of both techniques on the oral health-related quality of life for children and their families is another important area for future research.^21^ Understanding how each technique affects aspects such as pain, function, and overall well-being can provide valuable insights into the patient-centered outcomes of these treatments.

Further research is needed to comprehensively evaluate the long-term effects of the Hall Technique on occlusion and the eruption of permanent successor teeth.^16^ Longitudinal studies that monitor these aspects over time are necessary to address any potential concerns in these areas.

Finally, qualitative research exploring the perspectives and experiences of children, parents, and clinicians with both techniques could provide a richer understanding of their acceptability, feasibility, and perceived benefits and drawbacks.

### Implications for practice

Based on the results of this umbrella review and the quality of the evidence found, dentists who treat primary molars with healthy pulps or reversible pulpitis should consider that using HT in place of CT will be very unlikely to cause any added benefit or harm to children related to PMC success and survivability. Dentists should be reminded other outcomes of importance, such as treatment efficiency, cost-effectiveness, perceived child- and parent-acceptance, child discomfort, and child anxiety were not addressed in this present umbrella review. Hence, the dentist’s treatment options should be based upon clinical experience and the review of evidence-based publications.

Given the current evidence, pediatric dentists and general practitioners treating children should strongly consider the Hall Technique as a valuable and viable first-line treatment option for restoring carious primary molars, especially in situations where child cooperation is a concern or a less invasive approach is desired.^16^ The comparable success and survival rates, coupled with the added benefits of reduced treatment time and improved patient-parent acceptability, make the Hall Technique a compelling choice in many cases.^11,12^

The Conventional Crown technique remains a reliable and effective option, particularly in situations where complete caries removal is deemed clinically necessary or when dealing with specific case characteristics that might make the Hall Technique less suitable.^29^ The long-standing history and widespread familiarity with this technique also make it a comfortable and appropriate choice for many practitioners.

Clinicians should engage in open and informed discussions with parents regarding the available treatment options, clearly explaining the advantages and disadvantages of both the Hall Crown and Conventional Crown techniques. This should include addressing the aesthetic considerations associated with the preformed metal crowns used in the Hall Technique.^31^

Proper case selection is paramount for both techniques. The Hall Technique is specifically indicated for primary molars with carious lesions that do not show clinical or radiographic signs or symptoms of pulpal involvement.^29^ Thorough pulpal assessment is therefore a critical step before opting for the Hall Technique.

To ensure optimal patient care, clinicians should remain current with the latest research findings and consider incorporating the Hall Technique into their practice protocols and seeking continuing education opportunities to enhance their skills and knowledge in this area.^16^

### Implications for Dental Education and Policy

Dental school curricula should incorporate comprehensive and up-to-date training on the Hall Technique, ensuring that future dentists are proficient in both the Hall and Conventional methods of placing preformed metal crowns. This will equip new practitioners with the knowledge and skills to confidently offer the Hall Technique as a treatment option when appropriate.

Continuing education programs should offer opportunities for practicing dentists to learn about and implement the Hall Technique. Providing accessible and high-quality training can help to overcome any potential barriers to its adoption, such as a lack of familiarity or confidence with the technique.

Dental associations and guideline development committees should carefully consider the growing body of evidence supporting the Hall Technique when formulating clinical practice guidelines for pediatric preventive and restorative dentistry.^16^ Recognizing the Hall Technique as a valid and effective treatment option in relevant guidelines can promote its wider adoption in clinical practice.

By definition, HT is a secondary prevention intervention for arresting dental caries for primary molars, based upon the guidance from the American Academy of Pediatric Dentistry Reference Manual for restorative treatment and caries-risk assessment and management recommendations.^2,3^ This synthesis of the AAPD guidance supports the utilization of HT as a secondary prevention method^6–8^ for children with “moderate- to high-risk for dental caries, poor cooperation, or barriers to care.^3^”

## CONCLUSIONS

Based upon the results of this umbrella review, the following conclusions can be made:

1. There is no statistically significant difference for success and survival rates between the use of HT and CT in placing PMCs as demonstrated by comparable overall success and survival rates.
2. Dentists who treat primary molars with normal pulps or reversible pulpitis should consider, with moderate confidence, that treatment with HT in place of CT is highly unlikely to cause any added benefit or harm to children related to PMC success and survival.
3. The dentist’s treatment options for children requiring PMC treatment should be based upon clinical experience and the review of evidence-based literature.
4. Clinicians and policymakers should consider the AAPD guidance for restorative dentistry and caries-risk assessment and management for infants, children, and adolescents; which supports the utilization of HT as a secondary prevention method for arresting caries in children with “moderate- to high-risk for dental caries, poor cooperation, or barriers to care.”

## Data Availability

All data produced in the present work are contained in the manuscript.

## AKNOWLEDGMENTS

The authors extend appreciation and acknowledgment for funding to the Hansjorg Wyss Department of Plastic Surgery, NYU Langone Hospitals, New York, NY. The authors extend appreciation and acknowledgment for funding to the Hansjorg Wyss Department of Plastic Surgery, NYU Langone Hospitals, New York. This research was facilitated by Gemini Deep Research artificial intelligence (**AI**) tool (Google, Inc., Mountain View, CA), accessed on May 11, 2025, at the link, https://gemini.google.com/app. The authors acknowledge the tool’s assistance for the introduction section to identify key themes and for the discussion section to supplement the findings of other studies, which support or contradict the results, and enhance the implications of this umbrella review. The authors affirm that the original intent and meaning of the content remain unaltered during editing and that the AI tools had no involvement in shaping the intellectual content of this work. The authors assume full responsibility for upholding the integrity of the content presented in this manuscript.

## REFERENCES

1. Oral health: prevention is key. Lancet. 2009;373(9657):1. doi:10.1016/S0140-6736(08)61933-9

2. American Academy of Pediatric Dentistry. Pediatric restorative dentistry. The Reference Manual of Pediatric Dentistry. Chicago, Ill.: American Academy of Pediatric Dentistry; 2024:452-65.

3. American Academy of Pediatric Dentistry. Caries-risk assessment and management for infants, children, and adolescents. The Reference Manual of Pediatric Dentistry. Chicago, Ill.: American Academy of Pediatric Dentistry; 2024:306–12.

4. Badar SB, Tabassum S, Khan FR, Ghafoor R. Effectiveness of Hall Technique for Primary Carious Molars: A Systematic Review and Meta-analysis. Int J Clin Pediatr Dent. 2019;12(5):445–452. doi:10.5005/jp-journals-10005-1666

5. Lockerman LZ. Is the traditional placement of stainless steel crowns in primary teeth more painful than the non-preparation Hall technique? Evid Based Dent. 2023;24(3):108–109. doi:10.1038/s41432-023-00906-3

6. Kidd E, Fejerskov O, Nyvad B. Infected Dentine Revisited. Dent Update. 2015;42(9):802–809. doi:10.12968/denu.2015.42.9.802

7. Selwitz RH, Ismail AI, Pitts NB. Dental caries. Lancet. 2007;369(9555):51–59. doi:10.1016/S0140-6736(07)60031-2

8. Horst JA, Tanzer JM, Milgrom PM. Fluorides and Other Preventive Strategies for Tooth Decay. Dent Clin North Am. 2018;62(2):207–234. doi:10.1016/j.cden.2017.11.003

9. Hesse D, de Araujo MP, Olegário IC, Innes N, Raggio DP, Bonifácio CC. Atraumatic Restorative Treatment compared to the Hall Technique for occluso-proximal cavities in primary molars: study protocol for a randomized controlled trial. Trials. 2016;17:169. Published 2016 Mar 31. doi:10.1186/s13063-016-1270-z

10. BaniHani A, Santamaría RM, Hu S, Maden M, Albadri S. Minimal intervention dentistry for managing carious lesions into dentine in primary teeth: an umbrella review. Eur Arch Paediatr Dent. 2022;23(5):667–693. doi:10.1007/s40368-021-00675-6

11. Schwendicke F, Krois J, Robertson M, Splieth C, Santamaria R, Innes N. Cost-effectiveness of the Hall Technique in a Randomized Trial. J Dent Res. 2019;98(1):61–67. doi:10.1177/0022034518799742

12. Almaghrabi MA, Albadawi EA, Dahlan MA, Aljohani HR, Ahmed NM, Showlag RA. Exploring Parent’s Satisfaction and the Effectiveness of Preformed Metal Crowns Fitting by Hall Technique for Carious Primary Molars in Jeddah Region, Saudi Arabia: Findings of a Prospective Cohort Study. Patient Prefer Adherence. 2022;16:2497–2507. Published 2022 Sep 8. doi:10.2147/PPA.S370159

13. Stewart L, Moher D, Shekelle P. Why prospective registration of systematic reviews makes sense. Syst Rev. 2012;1:7. Published 2012 Feb 9. doi:10.1186/2046-4053-1-7

14. Page MJ, McKenzie JE, Bossuyt PM, et al. The PRISMA 2020 statement: an updated guideline for reporting systematic reviews. BMJ. 2021;372:n

15. Pollock M, Fernandes RM, Becker LA, Pieper D, Hartling L. Chapter V: Overviews of Reviews [last updated August 2023]. In: Higgins JPT, Thomas J, Chandler J, Cumpston M, Li T, Page MJ, Welch VA (editors). Cochrane Handbook for Systematic Reviews of Interventions version 6.5. Cochrane, 2024. Available from www.training.cochrane.org/handbook.

16. Chua DR, Tan BL, Nazzal H, Srinivasan N, Duggal MS, Tong HJ. Outcomes of preformed metal crowns placed with the conventional and Hall techniques: A systematic review and meta-analysis. Int J Paediatr Dent. 2023;33(2):141–157. doi:10.1111/ipd.13029

17. Pieper D, Antoine SL, Mathes T, Neugebauer EA, Eikermann M. Systematic review finds overlapping reviews were not mentioned in every other overview. J Clin Epidemiol. 2014;67(4):368–375. doi:10.1016/j.jclinepi.2013.11.007

18. Shea BJ, Reeves BC, Wells G, et al. AMSTAR 2: a critical appraisal tool for systematic reviews that include randomised or non-randomised studies of healthcare interventions, or both. BMJ. 2017;358:j4008. Published 2017 Sep 21. doi:10.1136/bmj.j4008

19. Higgins JP, Thompson SG, Deeks JJ, Altman DG. Measuring inconsistency in meta-analyses. BMJ. 2003;327(7414):557–560. doi:10.1136/bmj.327.7414.557

20. Bezerra CT, Grande AJ, Galvão VK, Santos DHMD, Atallah ÁN, Silva V. Assessment of the strength of recommendation and quality of evidence: GRADE checklist. A descriptive study. Sao Paulo Med J. 2022;140(6):829–836. doi:10.1590/1516-3180.2022.0043.R1.07042022

21. Hu S, BaniHani A, Nevitt S, Maden M, Santamaria RM, Albadri S. Hall technique for primary teeth: A systematic review and meta-analysis. Jpn Dent Sci Rev. 2022;58:286–297. doi:10.1016/j.jdsr.2022.09.003

22. Sterne JA, Sutton AJ, Ioannidis JP, et al. Recommendations for examining and interpreting funnel plot asymmetry in meta-analyses of randomised controlled trials. BMJ. 2011;343:d4002. Published 2011 Jul 22. doi:10.1136/bmj.d4002

23. Boyd DH, Thomson WM, Leon de la Barra S, et al. A Primary Care Randomized Controlled Trial of Hall and Conventional Restorative Techniques. JDR Clin Trans Res. 2021;6(2):205–212. doi:10.1177/2380084420933154

24. Elamin F, Abdelazeem N, Salah I, Mirghani Y, Wong F. A randomized clinical trial comparing Hall vs conventional technique in placing preformed metal crowns from Sudan. PLoS One. 2019; 14(6):e0217740. doi:10.1371/journal.pone.0217740

25. Ayedun O., Oredugba F., Sote E. Comparison of the treatment outcomes of the conventional stainless steel crown restorations and the hall technique in the treatment of carious primary molars. Niger J Clin Pr. 2021;24:584. doi: 10.4103/njcp.njcp_460_20.

26. Ebrahimi M., Shirazi A.S., Afshari E. Success and behavior during atraumatic restorative treatment, the Hall Technique, and the stainless steel crown technique for primary molar teeth. Pedia Dent. 2020;42:187–192.

27. Binladen H, Al Halabi M, Kowash M, Al Salami A, Khamis AH, Hussein I. A 24-month retrospective study of preformed metal crowns: the Hall technique versus the conventional preparation method. Eur Arch Paediatr Dent. 2021;22(1):67–75. doi:10.1007/s40368-020-00528-8

28. Ludwig KH, Fontana M, Vinson LA, Platt JA, Dean JA. The success of stainless steel crowns placed with the Hall technique: a retrospective study. J Am Dent Assoc. 2014;145(12):1248–1253. doi:10.14219/jada.2014.89

29. Şen Yavuz B, Kargul B. Comparison of the Hall Technique and Conventional Compomer Restorations: A 60-Month Follow-up. Clinical and Experimental Health Sciences. September 2023;13(3):541–548. doi:10.33808/clinexphealthsci.1105908

30. Tedesco TK, Innes NP, Gallegos CL, et al. Success rate of Hall Technique for restoring carious primary molars - systematic review and meta-analysis. Evid Based Dent. 2025;26(1):65–66. doi:10.1038/s41432-024-01044-0

31. Altoukhi DH, El-Housseiny AA. Hall Technique for Carious Primary Molars: A Review of Literature. Dentistry Journal. 2020; 8(1):11. 10.3390/dj8010011

